# Proteomics Investigation of Diverse Serological Patterns in COVID-19

**DOI:** 10.1101/2022.08.21.22278967

**Authors:** Xiao Liang, Rui Sun, Jing Wang, Kai Zhou, Jun Li, Shiyong Chen, Mengge Lyu, Sainan Li, Zhangzhi Xue, Yingqiu Shi, Yuting Xie, Qiushi Zhang, Xiao Yi, Juan Pan, Donglian Wang, Jiaqin Xu, Hongguo Zhu, Guangjun Zhu, Jiansheng Zhu, Yi Zhu, Yufen Zheng, Bo Shen, Tiannan Guo

**Affiliations:** Fudan University, Shanghai, China; Key Laboratory of Structural Biology of Zhejiang Province, School of Life Sciences, Westlake University, Hangzhou, Zhejiang, China; Center for Infectious Disease Research, Westlake Laboratory of Life Sciences and Biomedicine, Hangzhou, Zhejiang, China; Institute of Basic Medical Sciences, Westlake Institute for Advanced Study, Hangzhou, Zhejiang, China; Taizhou Hospital of Zhejiang Province affiliated to Wenzhou Medical University, Linhai, Zhejiang, China; Westlake Omics (Hangzhou) Biotechnology Co., Ltd., Hangzhou, Zhejiang, China

**Keywords:** COVID-19, serology, proteomics, inflammation, cellular immunity

## Abstract

Serum antibodies IgM and IgG are elevated during COVID-19 to defend against viral attack. Atypical results such as negative and abnormally high antibody expression were frequently observed whereas the underlying molecular mechanisms are elusive. In our cohort of 144 COVID-19 patients, 3.5% were both IgM and IgG negative whereas 29.2% remained only IgM negative. The remaining patients exhibited positive IgM and IgG expression, with 9.3% of them exhibiting over 20-fold higher titers of IgM than the others at their plateau. IgG titers in all of them were significantly boosted after vaccination in the second year. To investigate the underlying molecular mechanisms, we classed the patients into four groups with diverse serological patterns and analyzed their two-year clinical indicators. Additionally, we collected 111 serum samples for TMTpro-based longitudinal proteomic profiling and characterized 1494 proteins in total. We found that the continuously negative IgM and IgG expression during COVID-19 were associated with mild inflammatory reactions and high T cell responses. Low levels of serum IgD, inferior complement 1 activation of complement cascades, and insufficient cellular immune responses might collectively lead to compensatory serological responses, causing overexpression of IgM. Serum CD163 was positively correlated with antibody titers during seroconversion. This study suggests that patients with negative serology still developed cellular immunity for viral defense, and that high titers of IgM might not be favorable to COVID-19 recovery.

## Introduction

COVID-19 remains a threat to global health. The production of serum antibodies in the human body is a major defensive mechanism to neutralize SARS-CoV-2. Within them, IgM is initiated during the acute phase for early defense whereas IgG is secreted afterwards with a higher affinity for SARS-CoV-2 (1). Typically, COVID-19 patients underwent seroconversion (from negative to positive) of IgM and IgG within 20 days (2). The IgM and IgG expression kept elevating before reaching the plateau, with IgG plateau titers higher and longer-lasting than IgM plateau titers (3). The timespans of seroreversion (from positive to negative) were around 3-6 months since disease onset for IgM (4, 5) whereas hardly observed for IgG in one year (6). After vaccination, convalescent COVID-19 patients exhibited higher titers of IgM and IgG compared to healthy individuals (7).

Several atypical serological patterns were documented in the literature. 3.2% to 6.9% of the COVID-19 patients remained low expression or seronegative for both IgM and IgG throughout the disease stage (1, 2). It has also been reported that less than 10% of the patients exhibited 10-to 20-fold higher antibody titers than the average values when reaching the plateau (1, 8). These unexpected serological patterns indicate heterogeneous host responses during COVID-19, with unclear molecular mechanisms.

This study was designed to investigate the diverse expression patterns of IgM and IgG from a single-center cohort across two years of monitoring, and to explore the molecular evidence associated with atypical antibody expression via longitudinal proteomic profiling.

## Experimental procedures

### Ethics and data governance approval

This study has been approved by the Ethical/Institutional Review Board of Westlake University and Taizhou Hospital (approval notice: K20210218). The studies in this work abide by the Declaration of Helsinki principles. Since archived specimens were used, informed consent from the patients was waived by the boards. A flowchart of the study design is illustrated in Figure 1D.

**Figure 1.**
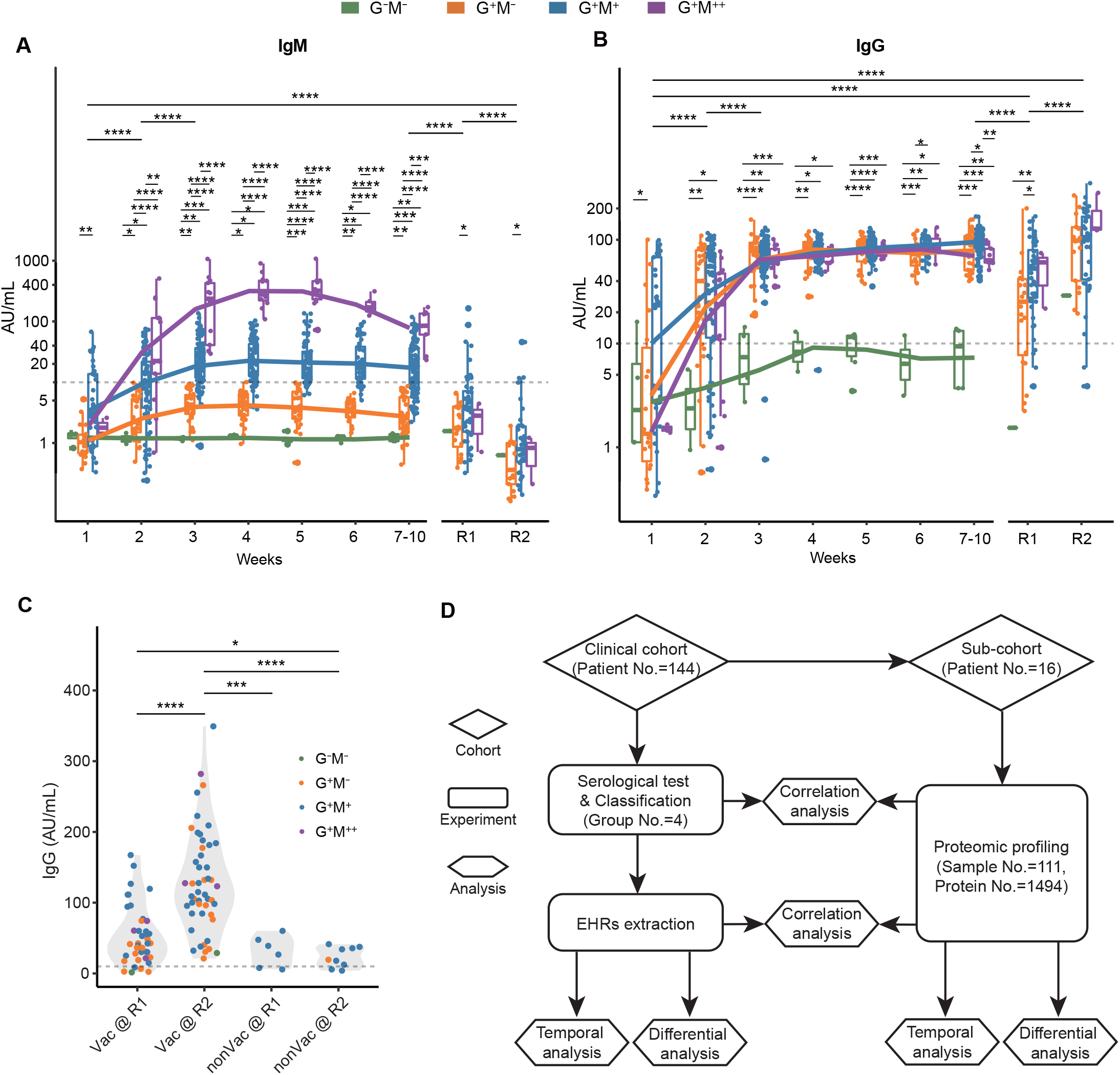
Overview of antibody expression in COVID-19. A-B) Classification and Two-year monitoring of SARS-CoV-2-specific IgM and IgG expression in COVID-19 patients. The y-axis denotes log-transformed antibody titers. The statistical significance was calculated within and across each timepoint. C) Comparison of IgG expression before and after vaccination. D) Study design for clinical and proteomic analyses of diverse serology in COVID-19.

### Patient information

We included 144 COVID-19 patients who were admitted to Taizhou Public Health Medical Center, Taizhou Hospital from January 17, 2020 to April 2, 2020. Within them, seventy-three patients participated in the one-year follow-up between day 363-397 (IQR, 10) since disease onset and fifty-eight patients participated in the two-year follow-up between day 728-763 (IQR, 7) since disease onset.

Information about demographics, epidemiological history, clinical symptoms, laboratory data, and hospitalization was collected through an electronic medical record system. All enrolled patients were confirmed to be infected with SARS-CoV-2 by use of real-time reverse-transcriptase polymerase-chain-reaction (RT-PCR) assay on the viral RNA extracted from nasopharyngeal or sputum specimens, and the classification of their disease severity were based on Diagnostic and Treatment Protocol for COVID-19 (Trial Version 5) issued by National Health Commission of the People’s Republic of China (9), unless otherwise mentioned. The onset date was defined as the day when any symptoms were noticed by the patients.

### Removal of identifying information

All information that would allow the patient/study participant or their family, friends or neighbors to identify them (e.g., age, past medical history, etc.) has been removed. Patient IDs from hospital records have been replaced with identifiers that cannot reveal the identity of the study subjects (e.g., R001, R002, etc.). The correspondence between identifiers and patient IDs was not known to anyone outside the research group.

### Laboratory characteristic tests

Nasopharyngeal or sputum specimens were collected to extract SARS-CoV-2 RNA, using a nucleic acid extractor (EX3600, Shanghai Zhijiang) and a virus nucleic acid extraction kit (P20200201, Shanghai Zhijiang). For nucleic acid detection, fluorescence quantitative PCR (ABI 7500, Thermo) coupled to a SARS-CoV-2 nucleic acid detection kit (P20200203, Shanghai Zhijiang) were used, which use a one-step RT-PCR combined with Taqman technology to detect RdRp, E, and N genes. “Positive” was concluded when the test for RdRp was positive (threshold cycle < 43) and one of the tests for E or N was positive (threshold cycle <43), or when two sequential tests of RdRp were positive whereas the tests for E and N were negative. Other laboratory characteristic tests were conducted according to the manufacturer’s protocols.

### Antibody analyses

The IgM and IgG antibodies against SARS-CoV-2 in serum samples were measured with the chemiluminescence immunoassay (CLIA) kits (iFLASH3000, Shenzhen YHLO). CLIAs were conducted based on two-step indirect immunization according to the manufacturer’s instructions. The recombinant nucleocapsid protein (N) and spike protein (S) antigens of SARS-CoV-2 were enveloped on magnetic beads, and an acridine ester labeled mouse anti-human IgM/IgG antibody was used as the detection antibody. The IgM/IgG antibody concentrations were positively correlated with the Relative Luminescence Unit (RLU). The cut-off to determine positivity was set at 10 AU/mL.

The neutralizing antibodies (NAbs) against SARS-CoV-2 in serum samples were measured using CLIA kits (Caris 200, Wantai). CLIAs for neutralizing antibody detection were based on competition immunization according to the manufacturer’s instructions. The neutralizing antibody in the sample and the biotinylated SARS-CoV-2 specific antibody compete with acridine ester labeled S protein. Next, streptavidin-coated magnetic particles were added. Through the interaction of biotin and streptavidin, a complex consisting of the magnetic particles coated by streptavidin, biotinylated SARS-CoV-2 specific antibody, and acridine ester S protein was formed. After washing and removing the substances that do not bind to the magnetic particles. The neutralizing antibody concentration in the sample was inversely proportional to the instrumentally detected RLU. The cut-off value to determine NAb positivity was set at 0.1 μg/mL.

### Patient classification based on antibody titers

The classification of COVID-19 patients was based on the maximum expression levels of IgM and IgG when reaching the plateau. The cut-off value, as determined by the detection kit, was 10 AU/mL. A total of 47 patients had IgM titers below the cut-off value (IgM^−^) and three had IgG titers below the cut-off value (IgG^−^) They were classified as seronegative for IgM and IgG, respectively.

Two patients had IgG titers above but close to the cut-off value. Specifically, the IgG plateau was reached on the second and the 18^th^ day after the symptoms’ onset for patients R055 (16.11 AU/mL) and R101 (13.80 AU/mL), respectively. These were the “outliers” of the patients with IgG plateau titers above the cut-off value, accordingly to Tukey’s test (Figure S2A). Clinicians classified them as seronegative for IgG (IgG ^−^)Similarly, nine patients with very high IgM expression were the “outliers” in the patients with IgM plateau titers above the cut-off value, accordingly to Tukey’s test. Clinicians classed them as patients with abnormally high IgM expression (IgM ^++^).

### Proteomic analysis

Serum samples were heated at 56 °C for over 60 min for inactivation. The sample preparation procedures were conducted as described previously (10) with several modifications: 10 μL serum from each specimen of the sample cohort was extracted and loaded onto High Select Top14 Abundant Protein Depletion Mini Spin Columns (Thermo Scientific) for high abundance protein depletion. Eluates were concentrated using Pierce™ Protein Concentrators PES, 3K MWCO (Thermo Scientific), and denatured with 50 μL lysis buffer (6 M urea and 2 M thiourea in 0.1 M triethylammonium bicarbonate, TEAB) at 31 °C for 30 min. The extracts were reduced with 10 mM tris (2-carboxyethyl) phosphine (Damas-beta) at 31°C for 40 min and alkylated with 40 mM iodoacetamide (Sigma) at 25°C for 40 min in darkness. The samples were diluted with 0.1 M TEAB buffer till the final concentration of urea was below 1.6 M, and trypsinized (Hualishi Technology) in double-step with an enzyme-to-substrate ratio of 1:20 at 31°C, for 60 min and 120 min, respectively. Trypsinization was stopped by adding trifluoroacetic acid (Damas-beta) till the final concentration of 1%, and digests were desalted with SOLAμ (Thermo Fisher Scientific), following the manufacturer’s instructions. Clean peptides were labeled with TMTpro 16plex label reagent sets (Thermo Scientific) according to the labeling set (Figure S2). A pooled sample was generated for labeling efficiency tests to ensure an incorporation ratio of over 95%. Afterward, samples in the batch were combined and fractionated in previously described settings (10). 60 fractions were derived and consolidated into 26 combined fractions. The fractions were dried and re-dissolved in 30 μL 2% ACN/0.1% formic acid. For nanoLC-MS/MS analysis, an EASY-nLC™ 1200 system (Thermo Fisher Scientific) coupled with Q Exactive HF-X hybrid Quadrupole-Orbitrap (Thermo Fisher Scientific) was applied, and data-dependent acquisition (DDA) mode was used throughout the analysis. The parameters were as previously described (10), except a 60 min LC gradient was applied for each acquisition. The resultant data were analyzed with Proteome Discoverer (Version 2.4.1.15). Protein database was a *Homo sapiens fasta* file downloaded from UniprotKB on April 10, 2020 and other parameter settings were as previously described (10).

### Statistical analysis

For clinical data (demographic information and clinical indicators), p values were calculated by two-sided Wilcoxon rank-sum tests. For proteomics data, p values were calculated by one-way analysis of variance (ANOVA) in the comparisons of divergent antibody expression and by two-sided Student’s t-test in other analyses. Benjamini & Hochberg correction was applied for p-value adjustment and labeled as *adjusted p*. Paired analyses were applied in comparing DEPs and in the comparisons of clinical indicators from organ dysfunction. Signs: *, p < 0.05; **, p < 0.01; ***, p < 0.005; ****, p < 0.001. Spearman correlation coefficients were calculated for the correlation of clinical data and proteomic data. LOESS (locally weighted scatterplot smoothing) model was applied for fitting analysis. Unsupervised hierarchical clustering was applied to display the DEPs from dysregulated seroconversion.

Categorical variables were described as frequency and percentage, and continuous variables were shown as mean and standard deviation or median and interquartile range (IQR) values as appropriate. To compare the continuous variables for data from different patient groups, an independent t-test was used when the data were normally distributed; otherwise, the Kruskal-Wallis H test was conducted. The categorical variables were compared using the χ^2^ test and Fisher’s exact test as appropriate. Statistical analyses were performed with R software (version 3.6.0).

### Pathway analysis

Metascape (11), String (12), and Ingenuine Pathway Analysis were applied for pathway enrichment in this study.

## Results

### Negative and exceptionally high IgM and IgG expression in COVID-19

We procured a cohort of 144 COVID-19 patients and used chemiluminescence immunoassays (CLIAs) to assess their expression levels of IgM and IgG (Methods). 790, 72, and 58 CLIA tests were conducted during the first 10 weeks since disease onset (weeks 1-10), the one-year revisit (R1), and the two-year revisit (R2), respectively. 84.5% of the revisited patients received vaccination between R1 and R2 (Table S1). Based on their plateau titers during weeks 1-10, the antibodies’ expression patterns were classified as follows: - for negative results or very low expression, for positive results, and for exceptionally high expression (Methods). Accordingly, patients were classified into four groups: IgG^−^ IgM^−^ (G ^−^M^−^, N = 5), IgG^+^ IgM^−^ (G^+^ M^−^, N = 42), IgG^+^ IgM^+^ (G^+^ M^+^, N = 88), and IgG^+^ IgM^+ +^ (G^+^ M^+ +^, N = 9) (Figure S1A and S1B).

We firstly assessed the IgM expression dynamics in the four groups. G^+^ M^+^ and G^+^ M^+ +^ patients underwent IgM seroconversion between weeks 1-2 (Figure 1A). However, the IgM titers in the G^+^ M^+ +^ group were significantly higher since week 2 and were over 20-fold in expression when reaching the plateau at week 4, compared to that in the G M group. The IgM titers in the G^+^ M^++^group remained over ten-fold higher than the G^+^ M^+^ group at weeks 7-10. Comparatively, none of the patients in the G^+^ M^−^or the G^−^ M^−^ group underwent IgM seroconversion during weeks 1-10. The overall IgM titers of the four groups had a significant decrease from weeks 1-10 to R1, which were further decreased at R2. Only 13.9% (N = 10) and 5.2% (N = 3) of the revisited patients were IgM seropositive at R1 and R2, respectively. The statistical differences in IgM titers between the G^+^ M^−^ and the G^+^ M^+^ groups persisted at R1 and R2, suggesting a long-term effect of COVID-19.

As for the IgG expression dynamics, all the groups except the G^−^ M^−^ group underwent seroconversion at weeks 1-2 and reached the plateau at week 3 (Figure 1B). Their IgG titers remained at plateau levels during weeks 3-6. Thereafter, the IgG titers exhibited a mild decrease in the G^+^ M^+ +^ group at weeks 7-10. By contrast, IgG seroconversion was hardly observed in the G^−^ M^−^ group, and most of the IgG titers in the G^−^ M^−^ group were below the positivity cut-off value during weeks 1-10. The overall IgG titers in the four groups exhibited a significant decrease from weeks 1-10 to R1. Statistical differences in IgG titers between G^+^ M^−^ and G^+^ M^+^ patients were also observed at R1. The IgG expression of all of the groups was significantly enhanced from R1 to R2, which might be attributed to vaccination (Table S1).

### Vaccination boosts IgG expression in COVID-19

To evaluate the effect of vaccination on IgG expression, we classed the convalescent patients at R2 into non-vaccinated (nonVac, N = 9) and vaccinated (Vac, N = 49) groups and compared their IgG titers (Figure 1C). The Vac group exhibited significantly elevated IgG titers from R1 to R2. All of the Vac patients were seropositive at R2, including one G^−^M^−^ patient R009. Comparatively, IgG titers in the nonVac group were equivalent between R1 and R2. These observations showed that vaccination can significantly boost IgG expression for COVID-19 convalescents. We also conducted 255 CLIA tests of neutralizing antibody (NAb) expression in 85 patients from the cohort throughout weeks 1-10 and R1 (Table S1). The NAb titers were relatively higher in the G^+^ M^+^ and G^+^M^+ +^ groups (Figure S1C) compared to others, suggestive of their stronger neutralizing abilities. NAb and IgG were positively correlated in all of the groups except for G^−^ M^−^ (Figure S1C), whereas NAb and IgM were not correlated (Figure S1D).

Our observation showed that COVID-19 patients underwent highly diverse antibody expression, especially IgM, after viral infection. IgM turned negative while IgG persisted with a significant decrease one year after COVID-19. IgG titers were significantly boosted after vaccination.

### COVID-19 severity and on-admission inflammation were positively correlated with antibody expression

We assessed the electronic hospital records (EHRs) of the enrolled patients (Figure 2 and Table S2). None of the G^−^M^−^ patients were severe cases, whereas 34.1% of the G^−^ M^−^ patients and 33.3% of the G^+^ M^++^ patients had severe symptoms, respectively (Figure 2A). Supportively, the G^+^ M ^+^and G^+^ M^+ +^ groups had significantly higher amounts of infected lung lobes and received more drug therapies, including immunoglobulin and methylprednisolone, compared to other groups (Table S2).

**Figure 2.**
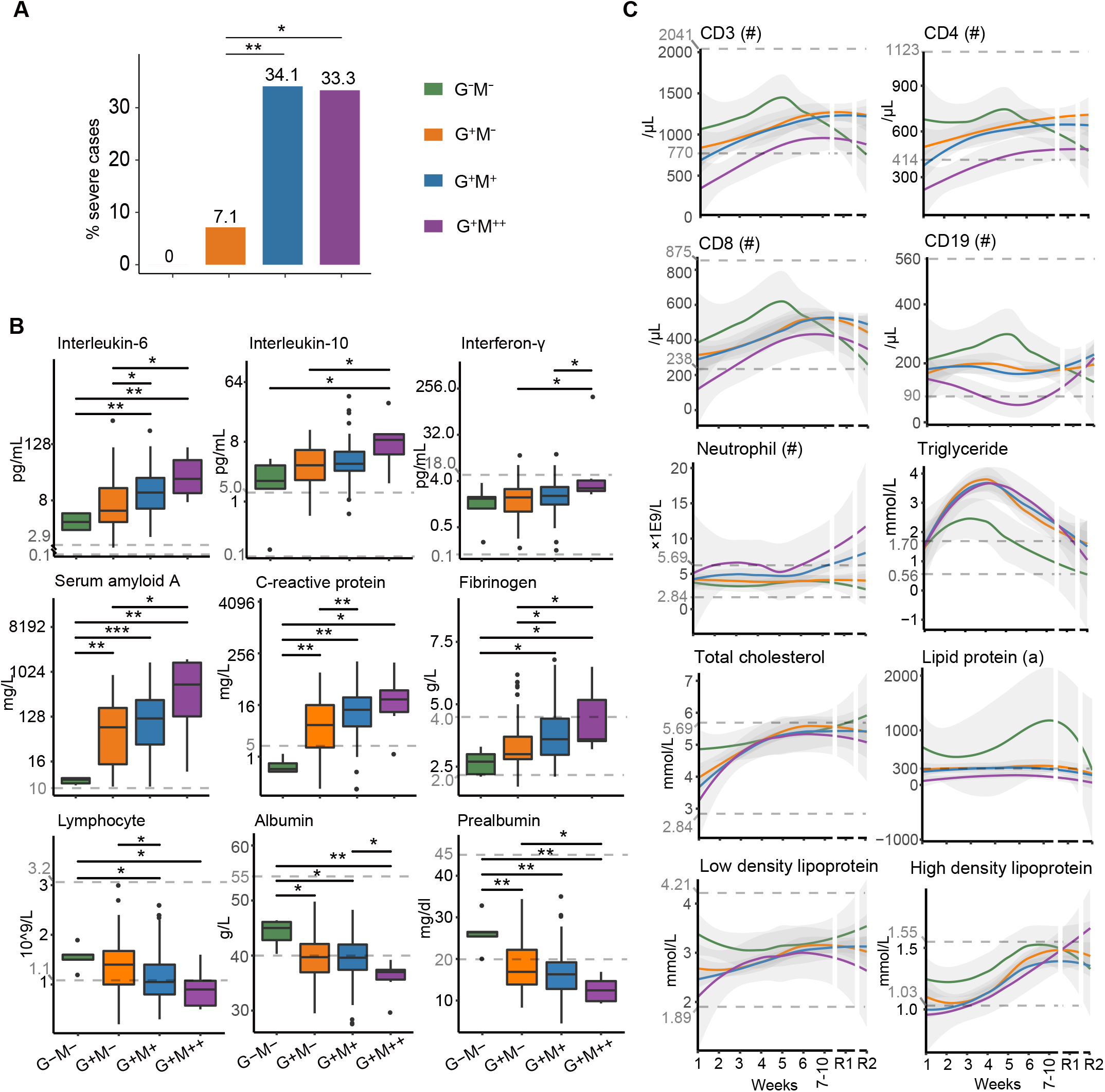
Clinical characteristics of patients with diverse serology. A) Distributions of severe cases in the four groups. B) Expression of nine selected clinical indicators on admission. C) Two-year temporal expression of ten selected clinical indicators.

To investigate the difference in the basic physiological status among the four groups, we compared their clinical indicators on admission (Figure 2B). Notably, risk factors such as SAA, CRP and inflammation factors such as IL-6, IL-10, and IFN-γ, were positively correlated with IgM and IgG expression, suggesting that patients with higher antibody titers also had stronger inflammatory responses. Coagulation factor fibrinogen remained at a normal range in the G^−^M^−^ group but overexpressed in some patients from the other groups, suggesting a tendency of coagulopathy in these patients. The lymphocyte amounts were negatively correlated with antibody titers and below the lower limit of normal ranges in most of the G^+^ M^++^ patients, suggesting that their baseline immunological status was inferior to the other patients. A series of nutritional factors such as albumin and pre-albumin, which have been reported as potential indicators for adverse outcomes in COVID-19 (13, 14), was negatively correlated with antibody expression titers.

To sum up, the severity and inflammation status of COVID-19 patients on admission were positively correlated with their antibody expression titers.

### Enhanced cellular immune responses are associated with negative IgM and IgG expression

We monitored the two-year temporal changes of serum clinical indicators in our cohort (Figure 2C). In the G M group, the expression of CD3, CD4, and CD8, three T lymphocyte markers, was below the lower limit of the reference ranges during weeks 1-3 and was significantly lower than the other groups during weeks 1-10. CD19, a B lymphocyte marker, was lower in the G^+^ M^+ +^ group than the other groups throughout weeks 1-10 and fell below the lower limit of the reference range since week 4. Comparatively, these CD markers were highly expressed in the G^−^ M^−^ group until week 5. The inter-group differences of these CD markers decreased after week 7.

These observations suggested that the G^−^ M^−^ patients exhibited stronger cellular immune responses compared to the other groups. Neutrophil counts during weeks 1-4 increased in the G^+^ M ^++^ group whereas remained equivalent in the other groups. Notably, starting from week 6, neutrophil counts in the G^+^ M^++^ and G^+^M^+^ groups increased beyond the upper limit of the reference range. The decrease of B and T lymphocytes and the increase of neutrophils during hospitalization have been previously observed in severe (15) and deceased (16) COVID-19 patients.

The temporal expression of several blood lipids and lipoproteins was different between the G^−^ M^−^ group and the others (Figure 2C). Triglyceride (TG) increased during weeks 1-3 followed by a gradual decrease in the four groups, but only the G^−^ M^−^ group went back to reference ranges by week 5. The G^−^M^−^ group also exhibited relatively higher expression of total cholesterol (TC), lipoprotein(a), low-density lipoprotein cholesterol (LDL-C), and high-density lipoprotein cholesterol (HDL-C) during weeks 1-10. The combination of low HDL-C and high TG, termed atherogenic dyslipidemia, was considered a risk factor for severe COVID-19 (17). These observations suggest different levels of systematic dyslipidemia during COVID-19, which were the mildest in the G^−^M^−^ group.

Taken together, patients who remained both IgM and IgG seronegative during disease had enhanced cellular immune responses whereas less perturbation of lipid metabolism, compared to the other patients.

### Serum proteomic investigation of four serological groups

To understand the differences in host responses behind the diverse antibody expression, we collected 111 serum samples from 16 representative COVID-19 patients (Figure S2A) to profile their longitudinal proteomic signatures during weeks 1-10 (Figure 3A). A total of 1494 proteins were characterized using TMTpro 16plex technology (Figure S2B and Table S3). The median value of the protein coefficient of variation (CV) for the pooled samples is 0.189 (Figure S2C), indicating high quality of our data.

**Figure 3.**
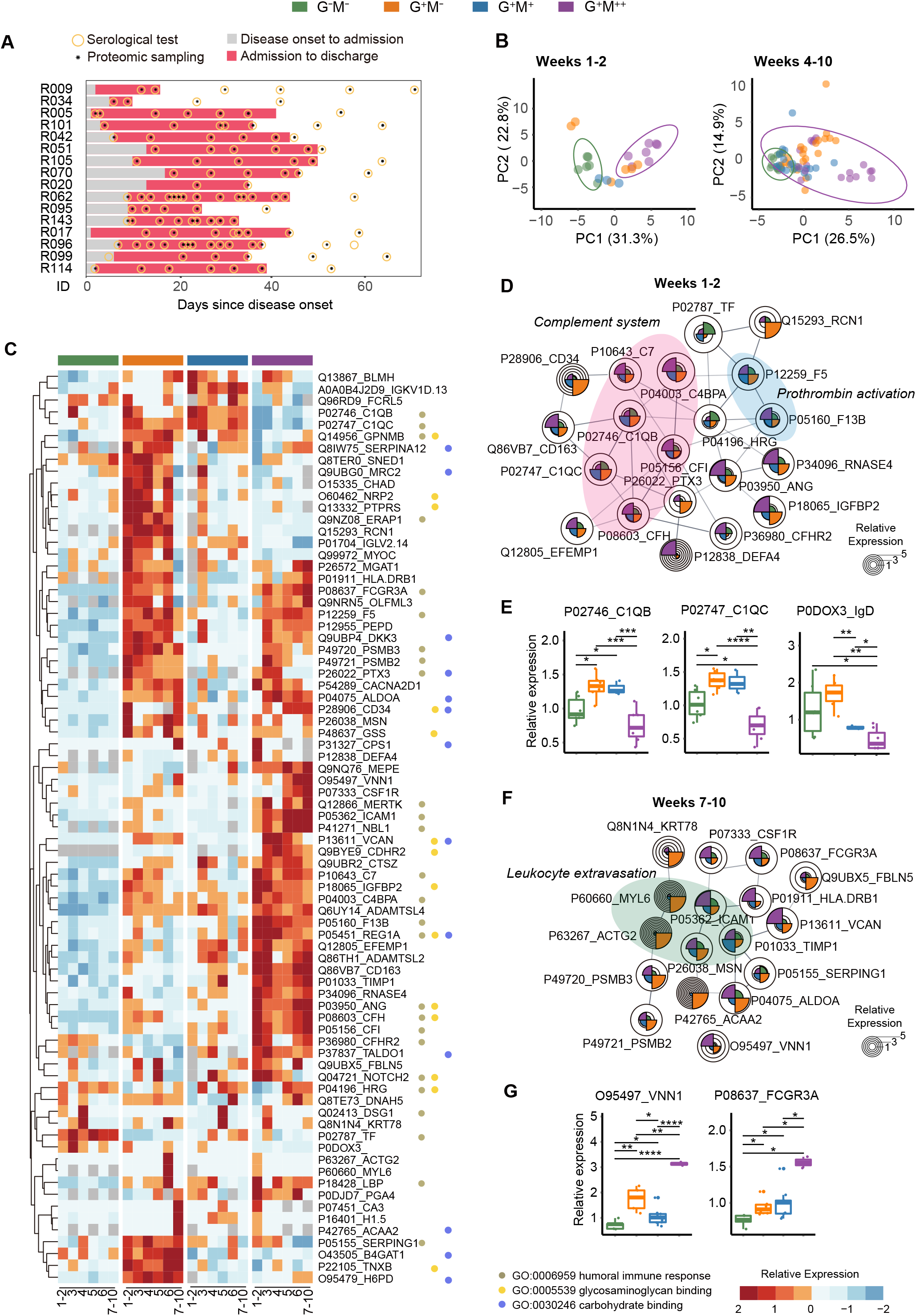
Longitudinal serum proteomics of 16 characteristic patients with diverse serology. A) Temporal serum sampling for the 16 characteristic patients. B) Heatmap and functional annotation of 77 DEPs across 10 weeks of proteomic profiling. C) PCA of serum samples stratified by the DEPs at weeks 1-2 and weeks 4-10, respectively. The ellipses are shown at a confidence interval of 95%. D) Interaction network of selected DEPs at weeks 1-2. E) Relative expression of DEPs C1QB, C1QC, and IgD at weeks 1-2. F) Interaction network of selected DEPs at weeks 7-10. G) Relative expression of DEPs VNN1 and FCGR3A at weeks 7-10.

To explore the proteomic differences among the four groups, ANOVA was used to assess differentially expressed proteins (DEPs) (adjusted p value < 0.05) at each week. 77 DEPs in total were identified, 61.0% (n = 47) of which were from weeks 1-2 (Figure S2D). Only one of these 47 proteins was overlapped with the DEPs from weeks 4-10, suggesting a distinct difference in the host responses during and after seroconversion. Accordingly, we found that the G^−^M^−^ and G^+^ M^++^ groups could be stratified by the DEPs during weeks 1-2 but not weeks 4-10 via principal component analysis (PCA, Figure 3B).

We next analyzed the key functions of the DEPs. The three most enriched pathways throughout weeks 1-10 are humoral immune responses, glycosaminoglycan binding, and carbohydrate binding (Figure 3C). Within them, humoral immune responses include multiple established risk factors of COVID-19, such as complement and fibrinogen factors (10, 18). The enrichment of Glycosaminoglycan binding might be attributed to either cytokine recognition or SARS-CoV-2 host entry (19). We then clustered the DEPs at the initial (weeks 1-2) and late (weeks 7-10) stages of COVID-19 via String (12). Complement system and prothrombin activation were the main enriched functions during weeks 1-2 (Figure 3D). Notably, the expression of C1QB and C1QC, subunits of complement 1 (C1), were significantly lower in the G^−^ M^−^ group compared to the G^+^ M^−^ and G^+^ M^+^ groups and further decreased in the G^+^ M^+ +^ group (Figure 3E). Comparatively, a list of downstream proteins of complement cascade, including C4BPA, C7, CFI, and CFH, were significantly upregulated in the G^+^M^+ +^ group (Figure 3D). This observation suggested that complement system was strongly activated in the G^+^M^+ +^ group but not via the classical C1-mediated pathway. P0DOX3, an IgD heavy chain residue, was negatively correlated with IgM expression in the four groups (Figure 3E), suggesting that IgD may compensate for the lack of IgM in the G^−^M^−^ and G^+^M^−^ groups (20). At weeks 7-10, leukocyte extravasation signaling was enriched (Figure 3F). Within them, VNN1, a proposed HDL regulator (21), was significantly upregulated in the G^+^M^++^ group (Figure 3G). FCGR3A was also overexpressed in the G M group, which might enhance the production of pro-inflammatory cytokines and the activities of cytotoxic effector cells (22).

In summary, the proteomic differences in the four groups were the most prominent during weeks 1-2, which were associated with complement cascades. The main host response differences during the late stage of COVID-19 relate to the leukocyte activities.

### Correlation between serum proteomes and clinical indicators

Beyond the functional analyses, we explored the correlation between the DEPs and the antibody titers during weeks 1-4 since disease onset. Five and four DEPs were significantly correlated with IgM and IgG expression titers (absolute value of Spearman correlation coefficient is over 0.5), respectively (Figure 4A). Within them, TIMP1, ICAM1, CD163, NOTCH2, and HLD-DRB1 are associated with inflammatory response, corroborating the important role of inflammation in serology. CD163, a marker for monocytic macrophages (23), is the most correlated protein with IgG among these regulators. As exemplified in Figure 4B, the expression of serum CD163 was positively correlated with antibody titers and was significantly higher than the other groups in the G M group throughout weeks 1-4, suggestive of their more activated macrophage polarization during COVID-19 (24).

**Figure 4.**
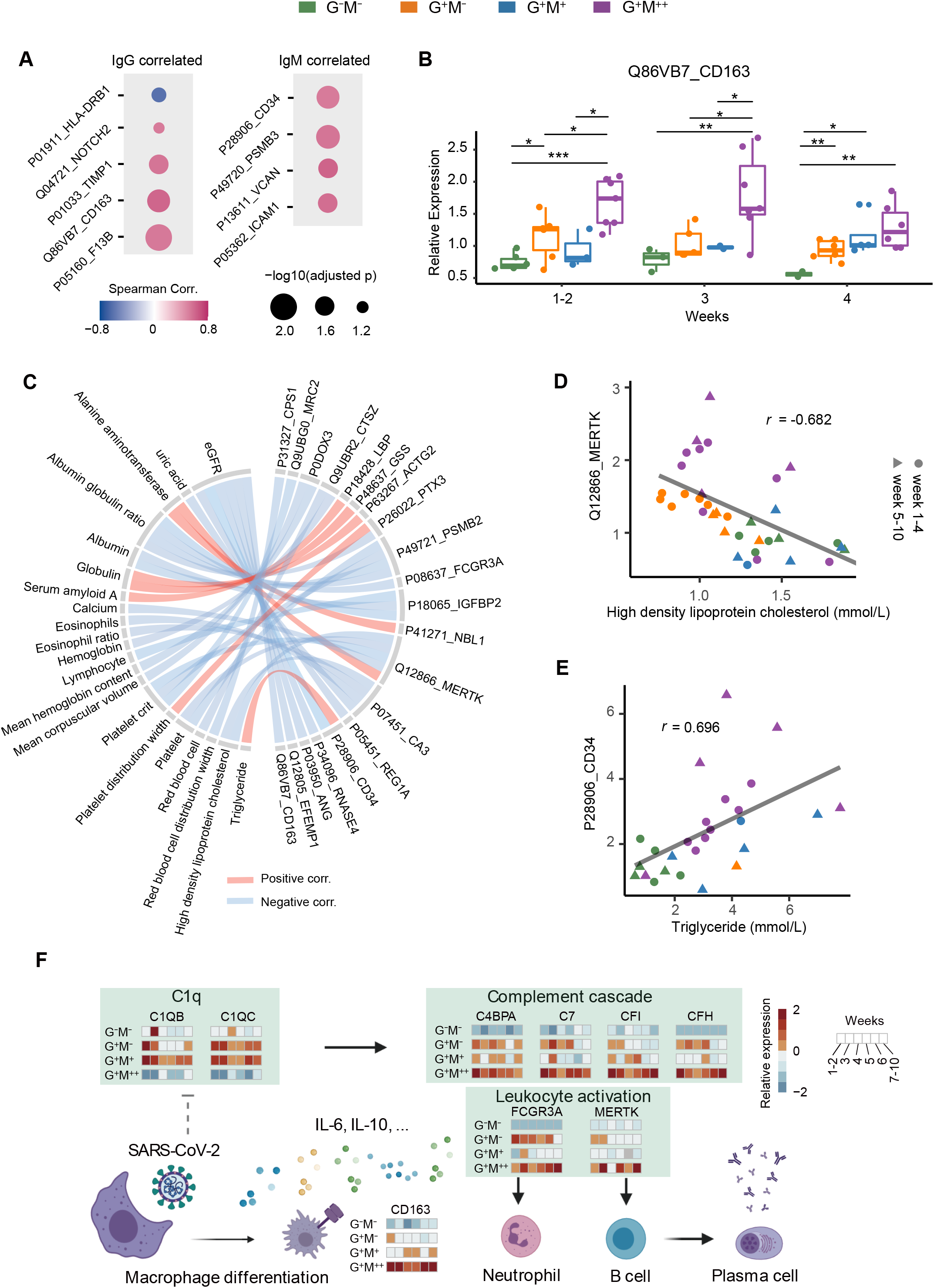
Correlations of DEPs with antibody and clinical indicators expression. A) Nine DEPs that were correlated with antibody expression during weeks 1-4 (absolute values of spearman correlation > 0.5). B) Temporal expression of CD163 during weeks 1-4. C) Correlations of clinical indicators with DEPs during weeks 1-10 (absolute values of spearman correlation > 0.65). D-E) Scatterplots of two sets of correlations: MERTK∼HDL-C and CD34∼TG, respectively. F) A putative working model for the host responses behind diverse COVID-19 serology.

We also assessed the correlation between clinical indicators and DEPs during weeks 1-10 (Figure 4C). HDL-C was significantly correlated with MERTK (Figure 4D), a transmembrane kinase that contributes to the B lymphocyte activation (25). TG was positively correlated with serum CD34 (Figure 4E), a human hematopoietic stem cell marker that also has a role in facilitating inflammatory cell trafficking. These evidences further suggested lipid involvement in the immunological activities during COVID-19.

### A putative working model for diverse serology in COVID-19

Based on our data, we propose a putative working model regarding the diverse COVID-19 serology (Figure 4F). Complement cascade is initiated upon disease onset, mediating the secretion of cytokines such as IL-6 and IL-10. This process could trigger macrophage polarization, as exemplified by the upregulation of CD163, which would further modulate local inflammation. The subsequently activated neutrophils and T lymphocytes could initiate B cell differentiation, leading to the activation of humoral immune responses to secret antibodies. These processes might be barely activated in the G^−^ M^−^ group due to *a prior* cellular immune responses that efficiently confront the invasion of SARS-CoV-2 upon disease onset. On the contrary, the cellular immunity might not be sufficient in the G^+^ M^++^ group for viral defense. Meanwhile, the activation of complement 1 might be disturbed in the G^+^M^++^ group. The high expression of inflammatory factors and rapidly ascending IgM titers might be a complementary process to defend against viral attacks.

## Discussion

In this study, we extracted two-year EHRs and applied TMTpro 16plex-based longitudinal proteomics to investigate the host responses of patients with diverse serology in COVID-19. We found that patients with negative IgM and IgG expression still developed strong T cell immunity for viral defense, and that the overexpression of IgM was associated with perturbed complement cascades and insufficient cellular immune responses.

Multiple studies have reported the association between antibody expression and COVID-19 severity. For example, Long *et al*. observed higher titers of IgM and IgG in severe patients than in mild patients since seroconversion (1). Garcia-Beltran *et al*. found that severe COVID-19 patients that required intubation or were passed away had the highest levels of IgG and IgA antibodies compared to others (26). However, none of them have studied the mechanisms underlying the differentiated antibody expression. Our manifestation that antibody expression was associated with on-admission inflammatory responses supports a previous speculation that severe disease might be caused by hyper-inflammation, which induces antibody overproduction (26). Conversely, high antibody titers might be involved in the antibody-dependent enhancement (ADE) of viral entry, which further induces the expression of inflammatory factors (27).

Our observation that a list of CD molecules (CD3, CD4, CD8, and CD19) were highly expressed in the G^−^M^−−^ patients whereas decreased in the G^+^M^++^ patients during COVID-19 suggested their differentially regulated lymphocytes to confront viral attacks. The low expression of T and B lymphocyte markers, namely lymphopenia, has been established as a severity hallmark in previous studies (15, 28), further supporting our data. Notably, intra-group differences of these molecules were not as significant after week 5, which might result from medical treatment (29). Therefore, the evaluation of lymphopenia on COVID-19 severity is recommended within one month since disease onset. Comparatively, CD163 as shown in our serum proteomics data was continuously upregulated during seroconversion in the G^+^M^+ +^ group. The high expression of CD163 has also been detected in the autopsy samples of six organs in the deceased COVID-19 patients (30), the peripheral blood mononuclear cells during COVID-19, and the THP-1 cell line after 48 hours of SARS-CoV-2 infection (24). A recent study reported that an overexpression of pulmonary CD163 might lead to idiopathic pulmonary fibrosis in COVID-19 (31). These observations collectively suggest that overly activated macrophage functions might not be favorable to COVID-19 recovery.

Although lipid metabolism dysregulation is typical in COVID-19 (10), our data showed that seronegative patients had the mildest symptoms of dyslipidemia compared to others. This might be attributed to their relatively milder inflammation, which mediates cytokine secretion that alters lipid homeostasis (32). We also found clues that HDL-C and TG might be associated with leukocyte activation during COVID-19. Mechanistic studies are needed in the future to explore their causality.

Our observation that SARS-CoV-2-specific IgG declined significantly one year after COVID-19 was in line with previous reports (33), suggesting a transition into immune memories. This also underlined the necessity of vaccinating recovered populations, which could significantly enhance IgG titers two years after COVID-19, according to our data. Notably, 85.7% (N = 42) of the vaccinated patients in our cohort received at least two doses of inactivated vaccines as of R2. Previous studies have shown that one to two doses of adenoviral vector or mRNA vaccines, were also viable to boost IgG and NAb titers in the recovered patients (7, 34). A longer-term serological monitoring of the same cohort is expected to understand the long-term humoral immunity in the vaccinated population with previous SARS-CoV-2 infection.

Our study has several limitations. Firstly, the single-center study with a relatively small patient cohort was possibly subject to demographic and experimental biases. Also, the sampling time points failed to cover the seroreversion stages for IgG. Furthermore, as the major purpose is to profile host responses associated with diverse serology patterns, we didn’t validate specific markers in this study.

## Data Availability

Patient information and serology data are available in the supplementary material. The proteomic raw data are deposited in ProteomeXchange Consortium (https://www.iprox.cn/page/PSV023.html;?url=1660551727880pH06, password: VcQ7).

## Abbreviations

ADE: antibody-dependent enhancement;
ARDS: acute respiratory distress syndrome;
C1: complement 1;
CLIA: chemiluminescence immunoassay;
COVID-19: Coronavirus Disease 2019;
CV: coefficient of variation;
DDA: data-dependent acquisition;
DEP: differentially expressed protein;
EHR: electronic hospital record;
HDL-C: high-density lipoprotein cholesterol;
IQR: interquartile range;
LDL-C: low-density lipoprotein cholesterol;
LOESS: locally weighted scatterplot smoothing Nab, neutralizing antibody;
nonVac: non-vaccinated;
PCA: principal component analysis;
R1: one-year revisit;
R2: two-year revisit;
RBD: receptor binding domain;
RLU: relative luminescence unit;
RT-PCR: reverse-transcriptase polymerase-chain-reaction;
TC: total cholesterol;
TEAB: triethylammonium bicarbonate;
TG: triglyceride;
TMT: tandem mass tag;
Vac: vaccinated.

## Acknowledgments

This work was supported by grants from National Key R&D Program of China (2021YFA1301602, 2020YFE0202200), National Natural Science Foundation of China (81972492, 21904107, 82072333), Zhejiang Provincial Natural Science Foundation for Distinguished Young Scholars (LR19C050001), Hangzhou Agriculture and Society Advancement Program (20190101A04), Medical Science and Technology Project of Zhejiang Province (2021KY394), Westlake Education Foundation, and Scientific Research Foundation of Taizhou Enze Medical Center (Group) (21EZZX01). We thank Westlake University Supercomputer Center for assistance in data storage and computation.

## Author Contributions

Y.Z., B.S., and T.G. designed the project, Y.Z., J.W., K.Z., D.W., and G.Z. collected the samples, X.L., J.W., K.Z., J.L, S.C., M.L., J.P., J.X., H.Z., and G.Z. organized the sample information, X.L., S.L., and X.Y. prepared the samples, X.L., M.L., Y.S., Y.X., and Q.Z. performed the data analysis, X.L., Y.X., and Q.Z. designed the figures, X.L. and R.S. wrote the manuscript, Y.Z., B.S., Y.Z., and T.G. supervised the project.

## Supplementary data

This article contains supplemental material Table S1-S3 and Figure S1-S2.

