## Supplementary material for "Proteomics Investigation of Diverse Serological Patterns in COVID-19": Figure S1; Figure S2

### Table of contents

Supplementary figure legends

Figure S1

Figure S2

Supplementary materials provided as separate files

### Supplementary figure legends

**Figure S1 Serology characteristics in COVID-19 patients.** A) Plateau expression of IgM and IgG labeled with different groups. B) Summary of patient and serology test number from different groups. C-D) Scatterplots of NAbs expression titers and their correlations with IgG and IgM expression titers, respectively.

**Figure S2 Proteomic experiment for the serum samples from the characteristic patients.** A) Temporal IgM and IgG expression of the 16 characteristic patients. B) Proteomic experimental workflow for serum sample preparation. C) Coefficient of variation for the pooled samples in eight batches. D) Summary of DEPs count from weeks 1-10.

### Figure S1

A

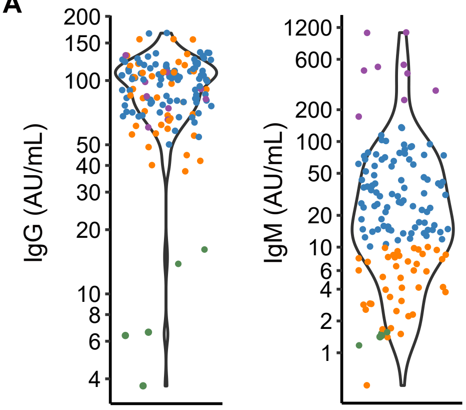


B

| Group | $G^-M^-$ | $G^+M^-$ | $G^+M^+$ | $G^+M^{++}$ |
|---|---|---|---|---|
| Patient No. | 5 | 42 | 88 | 9 |
| Serology test No. | 29 | 231 | 589 | 71 |

C

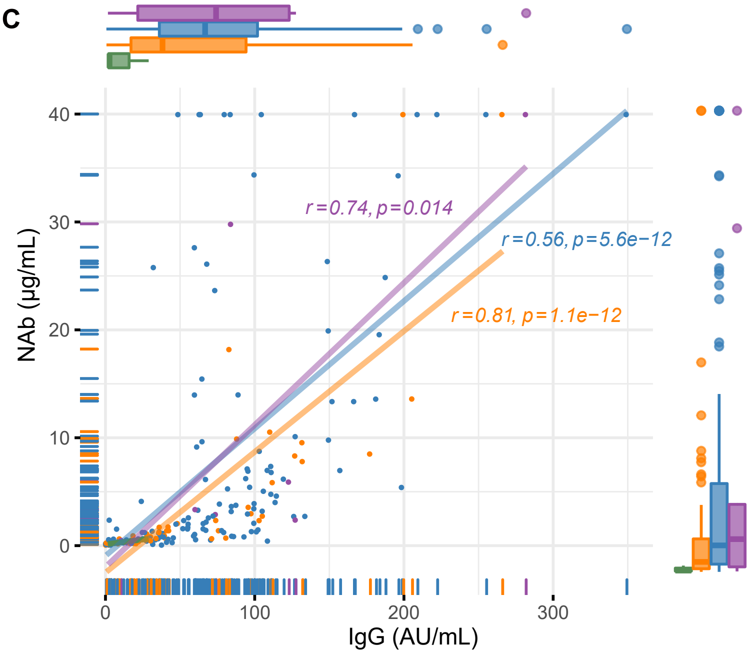


D

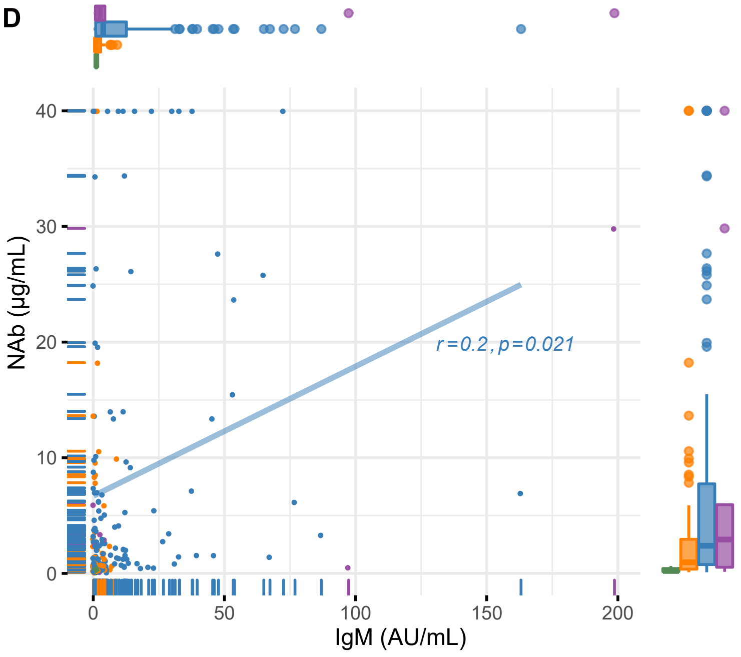

### Figure S2

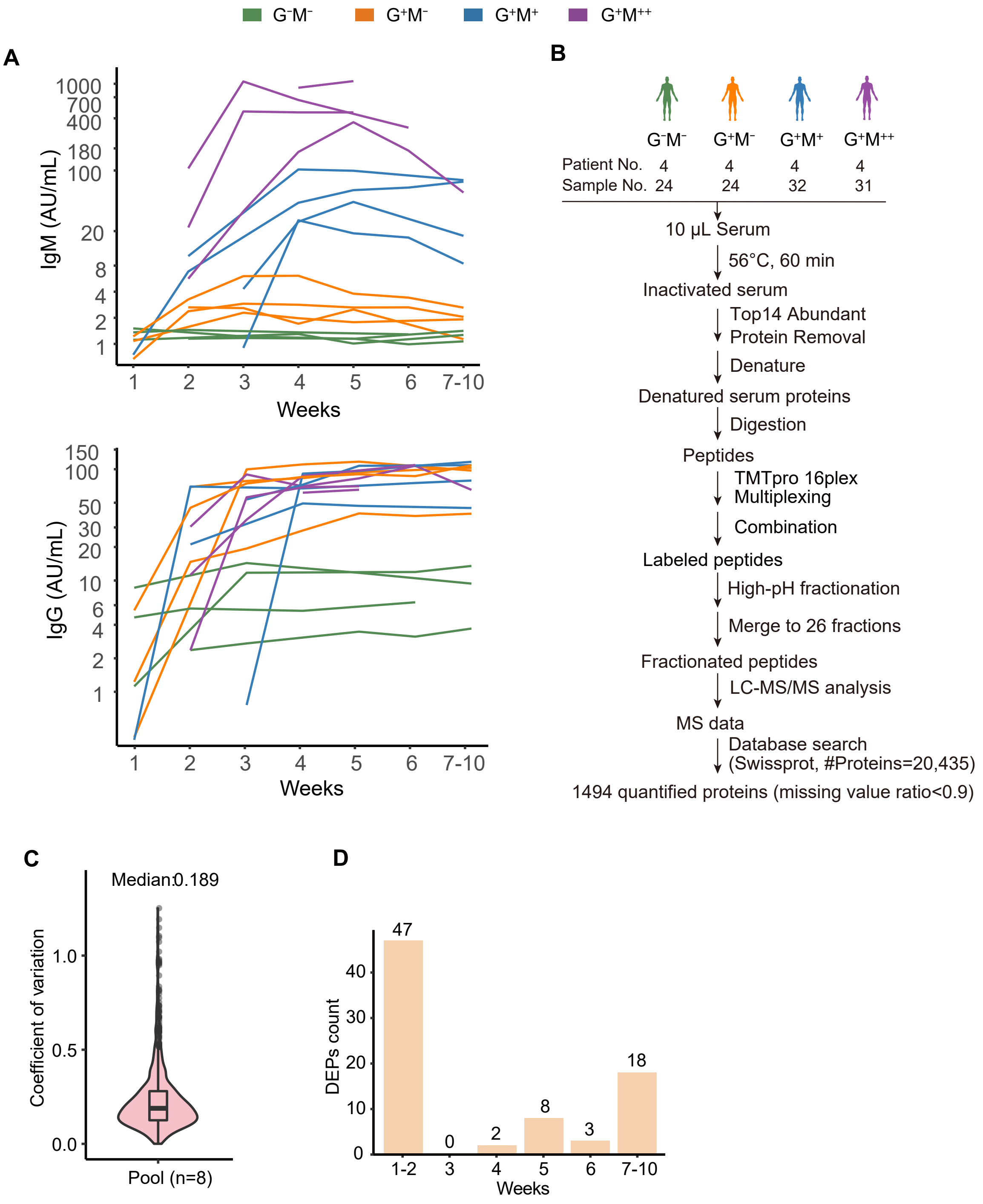
G⁻M⁻
G⁺M⁻
G⁺M⁺
G⁺M⁺⁺
A
IgM (AU/mL)
1000
700
400
180
100
20
8
4
2
1
1 2 3 4 5 6 7-10
Weeks
IgG (AU/mL)
150
100
50
30
20
10
6
4
2
1
1 2 3 4 5 6 7-10
Weeks
B
G⁻M⁻ G⁺M⁻ G⁺M⁺ G⁺M⁺⁺
Patient No. 4 4 4 4
Sample No. 24 24 32 31
10 μL Serum
56°C, 60 min
Inactivated serum
Top14 Abundant Protein Removal
Denature
Denatured serum proteins
Digestion
Peptides
TMTpro 16plex Multiplexing
Combination
Labeled peptides
High-pH fractionation
Merge to 26 fractions
Fractionated peptides
LC-MS/MS analysis
MS data
Database search (Swissprot, #Proteins=20,435)
1494 quantified proteins (missing value ratio<0.9)
C
Median:0.189
Coefficient of variation
1.0
0.5
0.0
Pool (n=8)
D
DEPs count
40
30
20
10
0
47
0
2
8
3
18
1-2 3 4 5 6 7-10
Weeks

### Supplementary materials provided as separate files

**Table S1. Basic information of the patient cohort.**

**Table S2 The comparison of clinical information of the patients with four categories of serology.** Categorical variables were expressed as numbers (percentages). The comparison of categorical variables between the four groups was performed using chi-square. The comparison of continuous variables between the four groups was performed using the Kruskal-Wallis statistics.

**Table S3. Proteomic results of the patient cohort.**
